# Clonal hematopoiesis in sickle cell disease

**DOI:** 10.1101/2021.06.12.21258772

**Authors:** L. Alexander Liggett, Liam D. Cato, Joshua S. Weinstock, Yingze Zhang, Seyed Mehdi Nouraie, Mark T. Gladwin, Melanie E. Garrett, Allison Ashley-Koch, Marilyn J. Telen, Brian Custer, Shannon Kelly, Carla Luana Dinardo, Ester C. Sabino, Paula Loureiro, Anna Bárbara Carneiro-Proietti, Cláudia Maximo, NHLBI Trans-Omics for Precision Medicine (TOPMed) Consortium, Alexander P. Reiner, Gonçalo R. Abecasis, David A. Williams, Pradeep Natarajan, Alexander G. Bick, Vijay G. Sankaran

**Affiliations:** Division of Hematology/Oncology, Boston Children’s Hospital and Department of Pediatric Oncology, Dana-Farber Cancer Institute, Harvard Medical School, Boston, MA, USA; Broad Institute of MIT and Harvard, Cambridge, MA, USA; Center for Statistical Genetics, Department of Biostatistics, University of Michigan School of Public Health, Ann Arbor, MI, USA; Department of Medicine, School of Medicine, University of Pittsburgh, Pittsburgh, PA, USA; Department of Medicine, Duke University Medical Center, Durham, NC, USA; Vitalant Research Institute, San Francisco, CA, USA; Department of Laboratory Medicine, University of California, San Francisco, San Francisco, CA, USA; Division of Pediatric Hematology, UCSF Benioff Children’s Hospital, Oakland, CA, USA; Fundação Pró-Sangue Hemocentro de São Paulo, São Paulo, Brazil; Institute of Tropical Medicine, Faculdade de Medicina da Universidade de São Paulo, São Paulo, Brazil; Pernambuco State Center of Hematology and Hemotherapy, Recife, Pernambuco, CEP 52011900, Brazil; Fundação Hemominas, Belo Horizonte, MG, Brazil; Fundação Hemorio, Rio de Janeiro, Brazil; Department of Epidemiology, University of Washington, Seattle, WA, USA; Division of Public Health Sciences, Fred Hutchinson Cancer Research Center, Seattle, WA, USA; Cardiovascular Research Center and Center for Genomic Medicine, Massachusetts General Hospital, Harvard Medical School, Boston, MA, USA; Division of Genetic Medicine, Department of Medicine, Vanderbilt University Medical Center, Nashville, TN, USA; Harvard Stem Cell Institute, Cambridge, MA 02138, USA

## Abstract

Curative gene therapies for sickle cell disease (SCD) are currently undergoing clinical evaluation. However, the occurrence of several myeloid malignancy cases in these trials has prompted safety concerns. Individuals with SCD are predisposed to myeloid malignancies, but the underlying causes remain undefined. Clonal hematopoiesis (CH) is a pre-malignant condition that also confers significant predisposition to myeloid cancers. While it has been speculated that CH may play a role in SCD-associated cancer predisposition, no data addressing this issue have been reported. Here, we leverage 74,190 whole-genome sequences to robustly study CH in SCD. While we have sufficient power to detect increased rates of CH, we find no variation in rate or clone properties between individuals affected by SCD and controls. These results should help guide ongoing efforts that seek to better define the risk factors underlying myeloid malignancy predisposition in SCD and help ensure that curative therapies can be more safely applied.

## 1. Main

In addition to the reduced life expectancy from disease complications [1], individuals with sickle cell disease (SCD) have a 3-10-fold increased life-time risk for acquiring acute myeloid leukemia (AML) and other myeloid malignancies [2, 3, 4]. These observations have recently generated profound interest due to a number of reports of myeloid malignancies arising during gene therapy that have halted several ongoing clinical trials [5, 6]. In one reported patient, the malignancy was not attributable to insertional mutagenesis, but the malignant clone was noted to harbor RUNX1, KRAS, and PTPN11 mutations [6]. Analysis of two SCD patients with myeloid malignancies in the setting of allogeneic transplantation revealed pre-existing hematopoietic clones with TP53 mutations that increased in size following conditioning, resulting in AML and myelodysplastic syndromes (MDS) [7]. However, the baseline rate of CH in SCD patients and thus the overall risk for myeloid malignancies in these individuals attributable to CH remains unclear. Myeloid malignancy predisposition is generally present in the setting of CH with a variant allele frequency (VAF) above 10% [8, 9, 10, 11, 12]. Here, we have robustly assessed the prevalence of CH in individuals with SCD who were subject to sequencing and variant calling in tandem with other unaffected individuals as part of the NHLBI Trans-Omics for Precision Medicine (TOPMed) consortium to interrogate whether SCD may predispose to CH [13].

To establish the prevalence of CH across human age, WGS data from 71,100 individuals unaffected by SCD and 3,090 individuals affected by SCD with ages of 70 years old and below were compiled from 30 distinct cohorts sequenced as part of the TOPMed consortium (Supplementary Figure 1). Previously described methods were employed to identify somatic variants and the presence of clonal expansions indicative of CH [14]. In short, somatic mutations in the blood were identified by GATK Mutect2 using a ‘panel of normals’ to correct for genomic loci that were hotspots for sequencing artifacts, followed by filtering of identified loci to those that are reliably classified as pre-leukemic driver mutations [15]. Samples were sequenced to an average depth of 38X, a depth that we previously showed was sufficient to reliably detect the majority of somatic mutations that exist at VAFs above 5% [14].

**Supplementary Figure 1:**
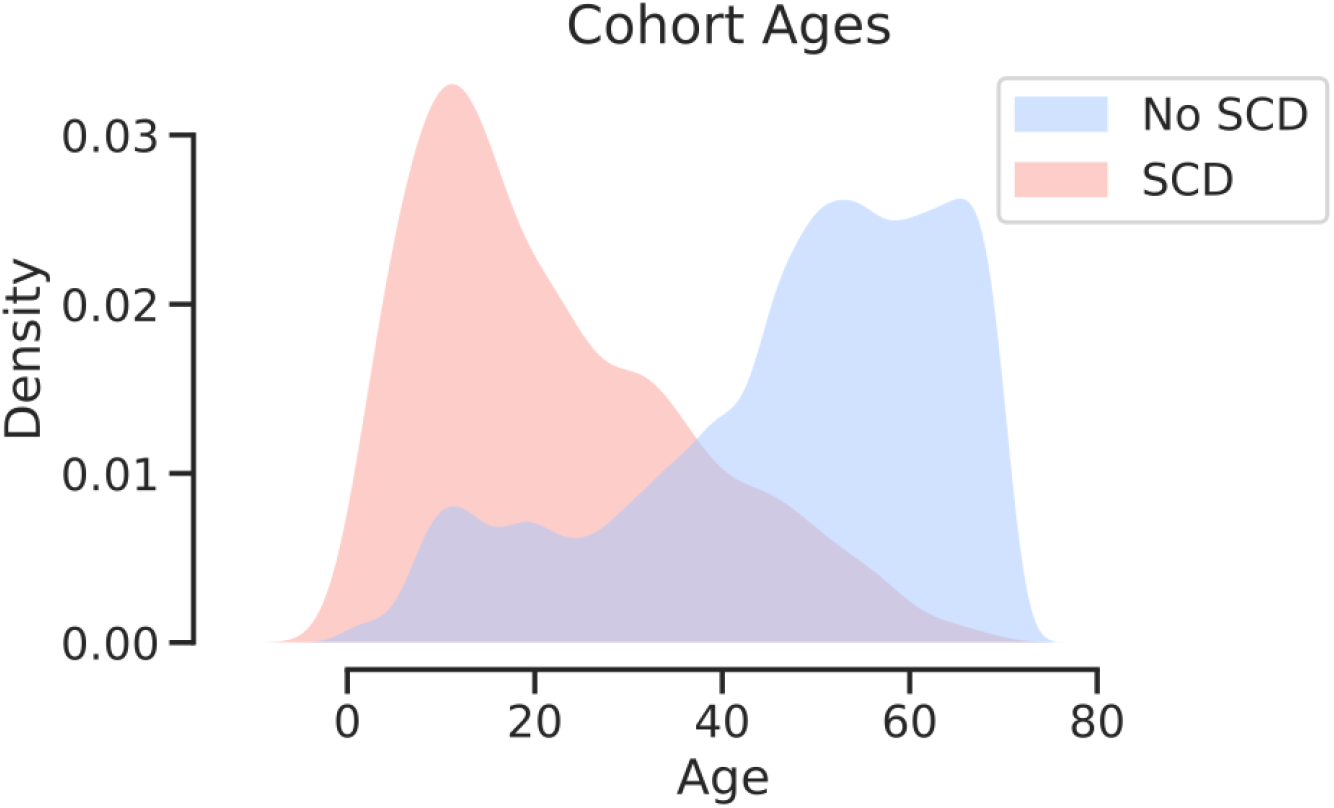
Cohort age distributions. Age distributions of all individuals combined from each of the TOPMed cohorts used in this study, separated by SCD status into those without SCD (blue) and those with SCD (red).

We identified 27 SCD patients and 3,063 unaffected individuals with CH. The prevalence of CH in the SCD cohorts and unaffected individuals both strongly correlated with age. There was, however, no significant difference in the prevalence of CH within the SCD cohort when compared to unaffected controls in unadjusted analyses (OR=1.30, p=0.20) (Figure 1A). Using a generalized linear model to account for covariates that are likely to impact the predicted CH prevalence, including age, age^2^, sex, study, and 10 principal components of genetic ancestry (PCs), we found that the prevalence of CH in individuals with SCD is not elevated over unaffected controls (SCD OR=1.55, p=0.20, Supplementary Figure 2). As a further sensitivity analysis, we performed a 1:20 case/control matching on age, sex, and the first ten PCs, as well as excluding several smaller disease-specific cohorts (detail in Methods), to identify a group of controls that were as similar as possible to our SCD cases. Even with this constructed set of unaffected controls, the CH prevalence in the SCD cohort was not significantly elevated, and even trended lower (OR=0.80, p=0.29, Figure 1B).

**Figure 1:**
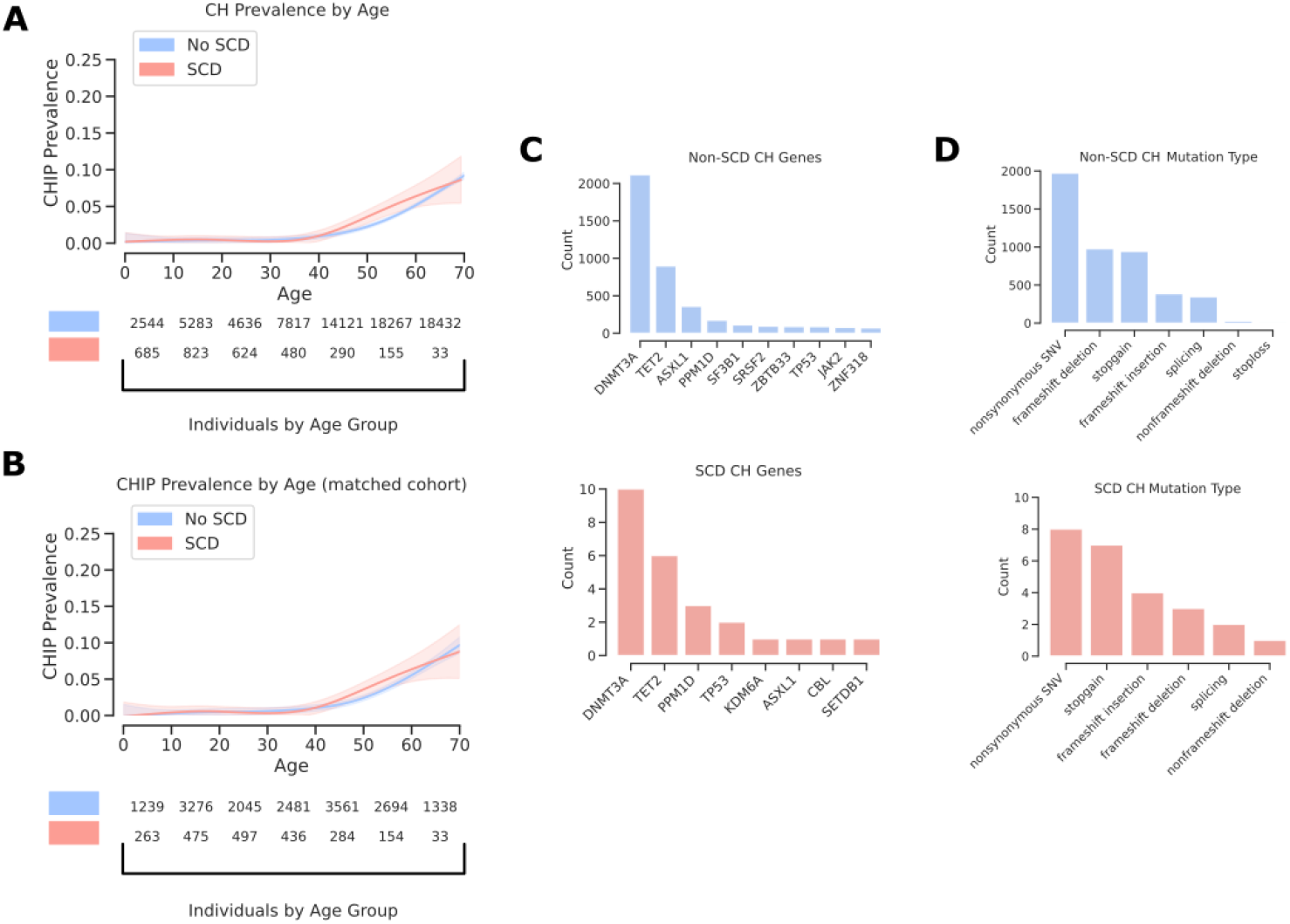
Prevalence of CH is similar in unaffected and SCD populations. A, A generalized additive model is used here to fit rates of CH within WGS data from a total of 71,100 individuals unaffected by SCD and 3,090 individuals affected by SCD, which indicates no significantly increased prevalence of CH within individuals affected by SCD (OR=1.30, p=0.20). B, the logistic regression model for effect estimates a is adjusted for age, age^2^, sex, study, and the first 10 principal components. C, Genes ranked by variant load across all individuals separated by SCD status into unaffected (blue) and affected (red). D, Type of genetic change ranked by prevalence in all individuals separated by SCD status into unaffected (blue) and affected (red).

**Supplementary Figure 2:**
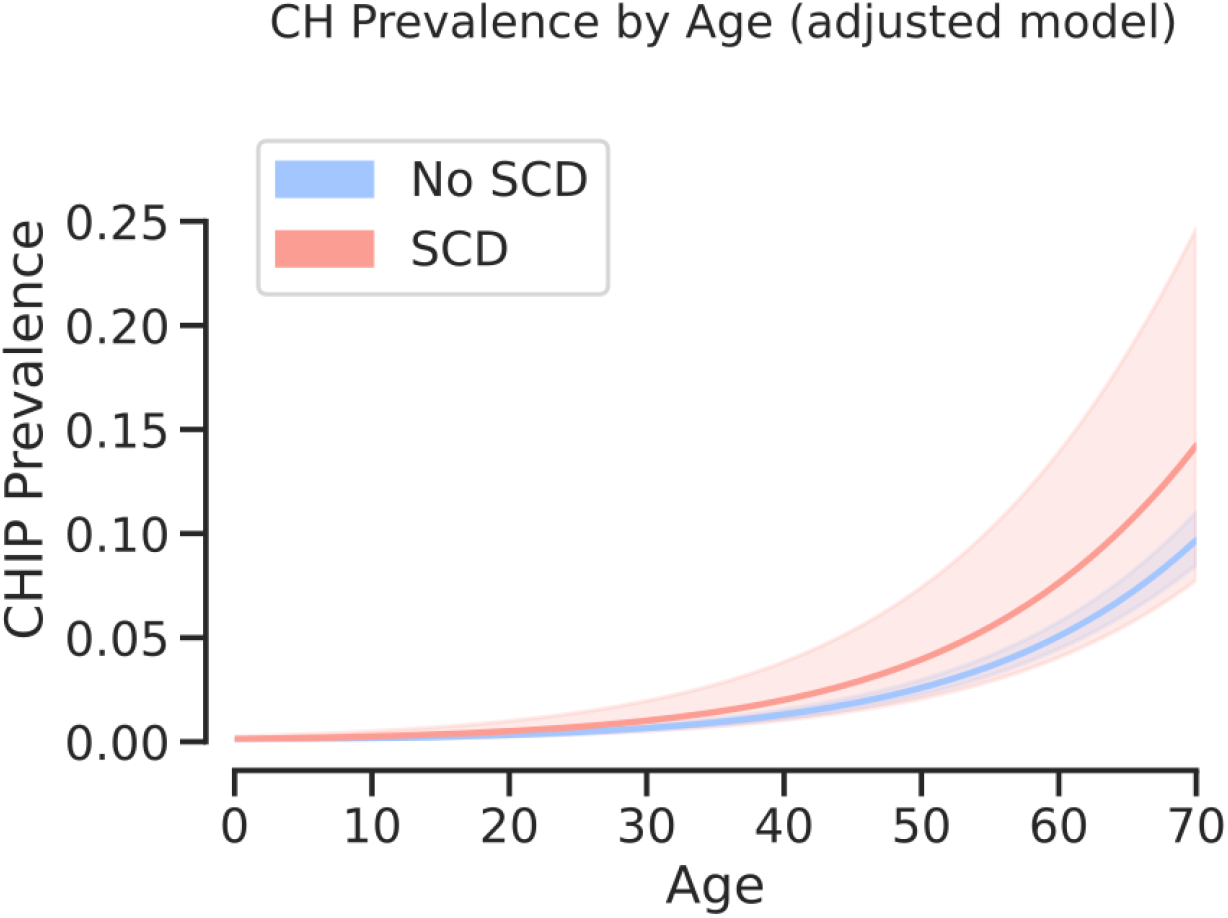
Matched background displays similar prevalence of CH as SCD cohort. 20 matched controls for each individual with SCD by matching the first 10 PCs, age, and sex, were pooled together to create a tailored unaffected cohort to compare to SCD affected individuals (OR=0.74, p=0.155).

Although our set of controls included individuals with sickle cell trait (2.6% of controls), we do not observe any altered rate of CH in these individuals compared to the other controls (OR=1.09, p=0.54) (Supplementary Figure 3). Similar subsetting of the SCD cohort to just those individuals with a hemoglobin SS (Hb SS) homozygous genotype, which is the most common SCD genotype, showed no significant difference in prevalence of CH between SS SCD and unaffected individuals (OR=1.34, p=0.22, Supplementary Figure 4). Furthermore, separating the different hemoglobin genotypes revealed that Hb S*β*0thalassemia was closest to a significantly different odds of CH, although it did not reach significance and only 84 individuals had this genotype (OR=3.81, p=0.067). Moreover, Hb S*β*+thalassemia (OR=1.21, p=0.85) and Hb SC disease (OR=1.02, p=0.97) were both far from significantly different with respect to non-SCD participants. Due unfortunately in part to the decreased survival in individuals with SCD, the cohort size diminished with age, but nonetheless with the cohorts available to us, we have 95% power to detect a 2-fold difference and 80% power to detect a 1.75-fold difference in the prevalence of CH in SCD.

**Supplementary Figure 4:**
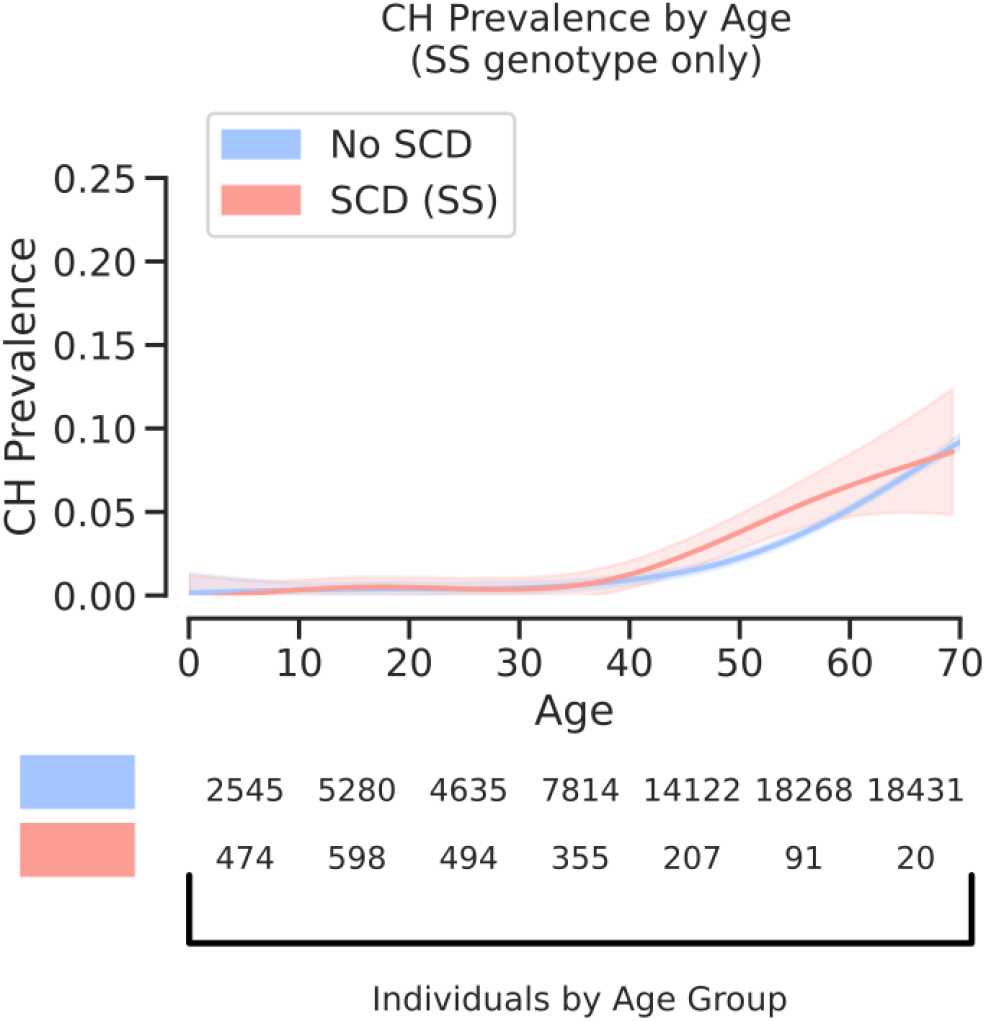
CH prevalence in SS SCD subgroup. Homozygous SS SCD genotypes alone do not show elevated prevalence of CH compared with unaffected individuals (OR=1.34, p=0.22).

Tissue microenvironmental and intrinsic conditions have a profound impact on the selective advantages and disadvantages conferred on cells by somatic mutations [16, 17]. We therefore expected that while the overall prevalence of CH in SCD may not be elevated, the oncogenic drivers of CH in SCD may notably differ. While categorizing somatic drivers of CH by affected gene and ranking by most prevalent results in slightly different spectra, no genes are significantly more likely to be mutated and represented in expanded clones in the context of SCD (Figure 1C). Similarly, classifying somatic mutations by type of change, exposes no significant differences in the types of somatic mutations that occur in the context of SCD (Figure 1D).

**Supplementary Figure 3:**
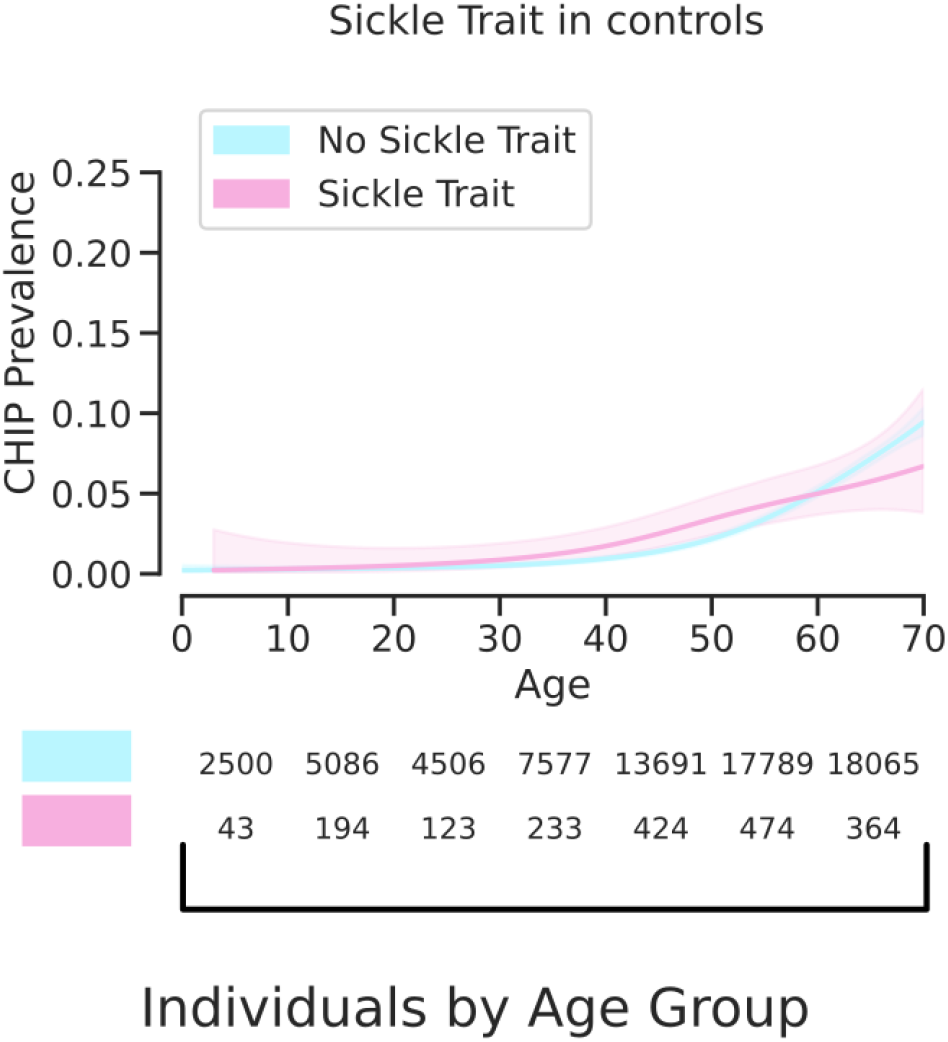
CH prevalence when accounting for sickle cell trait. From the group of all control individuals, those individuals who were heterozygous for the sickle cell trait (rs334) SNP are classified as having the sickle cell trait (pink) and those without it are classified as having no sickle cell trait (blue). Even when accounting for the rs334 SNP, there is no significant difference between the two groups.

By treating the number of CH variants per individual as an ordinal variable and correcting for age in a proportional odds logistic regression, having SCD did not correlate with an increased number of detected mutation-harboring clones (Supplementary Figure 5). The overwhelming majority (*>*70%) of individuals with CH in our dataset had only a single detectable somatic CH mutation, and this did not differ between individuals with SCD and unaffected individuals.

**Supplementary Figure 5:**
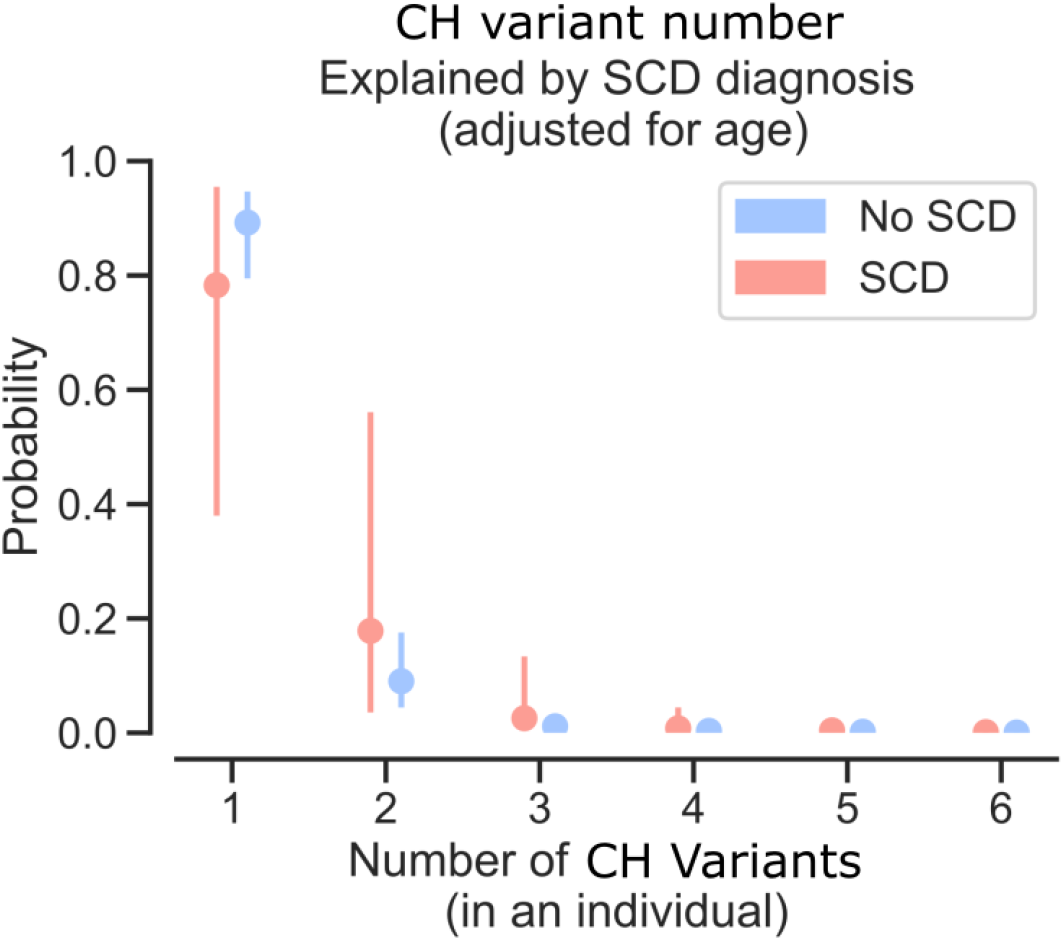
CH clone load per individual is unchanged by SCD. Number of variants per individual classified as indicative of CH appear at similar rates in unaffected individuals and individuals with SCD.

While overall rates of CH may not significantly differ among individuals with or without SCD, mutagenic processes may be altered by specific pathogenic features of SCD. We expect that such mutagenic processes could be reflected as alterations to the relative rates of single base substitutions (SBS). Importantly, the trinucleotide SBS mutation spectrum for the unaffected individuals showed no significant difference from those individuals with SCD (cosine similarity of 0.98, Figure 2A, B). This similarity suggests that at least as seen through SBS, there are no prominent mutagenic processes that are uniquely observed in the context of SCD.

**Figure 2:**
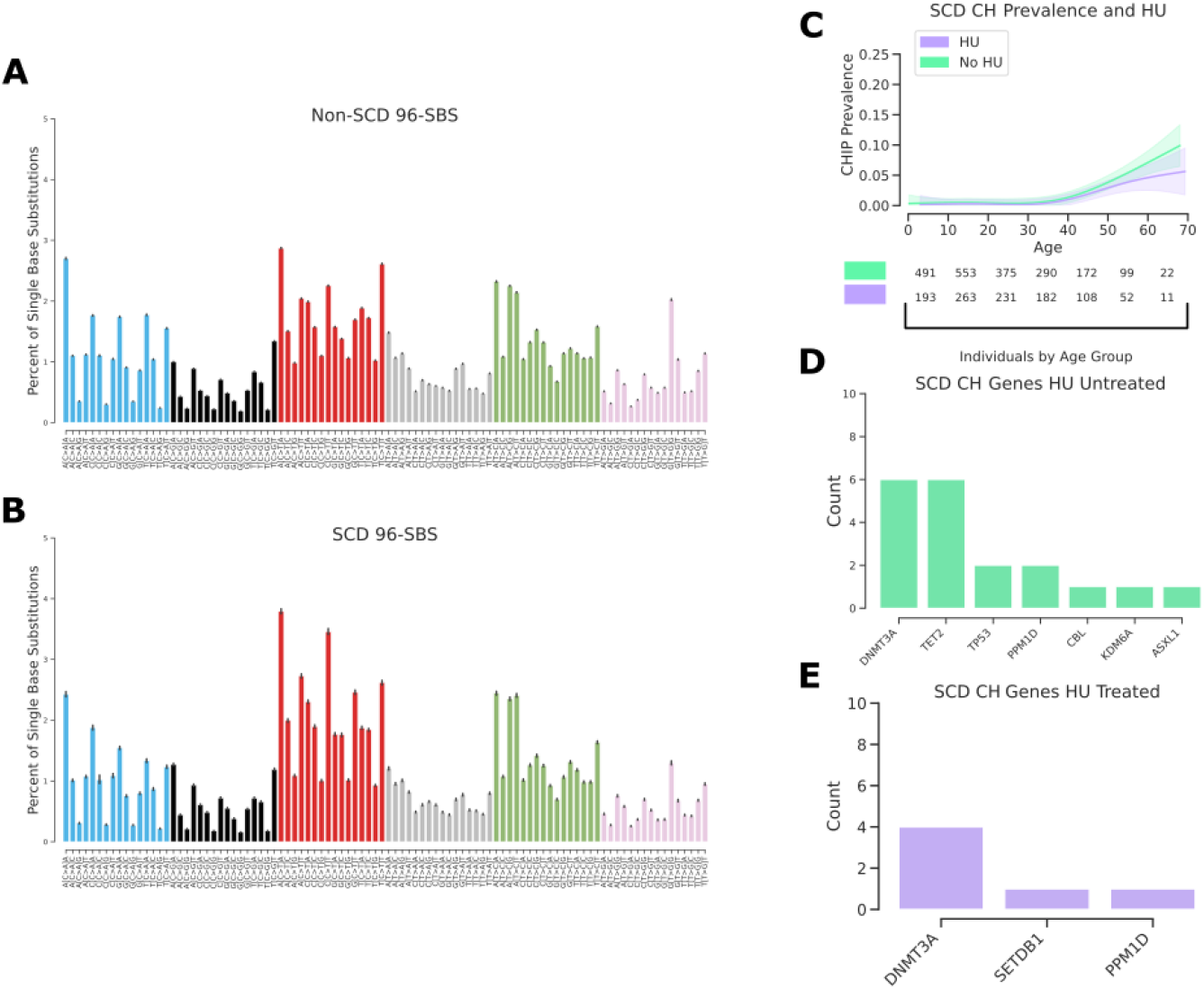
Hydroxyurea treatment does not impact the rate of CH development. Percent of total single-base substitutions made up by each possible 96 substitution and trinucleotide context pairs per individual (error is standard deviation across individuals). Signatures are separated into A, individuals without SCD and B, individuals with SCD. C, A generalized additive model is used here to fit rates of CH within WGS from individuals with SCD separated into never HU treated or HU treated groups. There is no significant difference in the rate of CH in either the HU treated or untreated groups adjusted for age, age^2^, sex, study, and the first 10 principal components (OR=0.58, p=0.23). D, Genes ranked by variant load across all individuals with SCD separated by HU treatment status into untreated (green) and D, treated (purple).

Hydroxyurea (HU) has been shown to have a significantly positive clinical impact in individuals with SCD and is thought to primarily influence disease severity by increasing levels of fetal hemoglobin (HbF) [18]. Given its cytotoxic effects and evidence from animal models, it has been posited that HU may be mutagenic and potentially carcinogenic, despite demonstrated safety profiles in vivo [19]. We therefore reasoned that while the overall SCD cohort may not exhibit an elevated risk of developing CH, those treated with HU may. Treating individuals with SCD categorically as either undergoing HU treatment or not, we found no significant elevation in CH prevalence in the group treated with HU (OR=0.58, p=0.23, Figure 2C). We additionally reasoned that while HU treatment may not drive an apparent increased prevalence of CH, it could select for cells with mutations in a different spectrum of genes. As before, we saw no significant difference in the spectrum of mutated genes in the HU-treated group (Figure 2D, E). There appeared to be a trend toward a reduced rate of CH in the HU treated individuals with SCD, but there was no significant difference in individuals over 50 years of age (OR=0.61, p=0.55).

Although not well studied, SCD has been associated in multiple studies with predisposition to myeloid malignancies and this has become of acute interest as the cancers observed in ongoing gene therapy trials have halted several current efforts [4]. CH is an important factor predisposing to myeloid malignancies and it has been speculated that individuals with SCD may have elevated rates of CH. We have taken advantage of the largest genome sequencing study of individuals with SCD to date and do not observe any increased rate of detectable CH. While our study is limited by the size of the SCD cohorts available, we would note that we are well powered to detect a 2-fold increased prevalence of CH with our current cohorts. Additionally, we detect CH using WGS and are unable to detect extremely small clones. However, the clinical implications of small clones in predisposing to myeloid malignancies is unknown and existing literature suggests that only larger clones with VAFs *>* 10% have prognostic impact [8, 9, 11, 10, 11]. While further studies are needed, these findings are likely to inform clinical decisions and suggest that CH surveillance as part of routine clinical care for SCD patients is no more indicated at the current time than it is in the general population. In the setting of clinical trials testing curative gene therapy or genome editing approaches, CH surveillance may not provide an effective approach to mitigate malignancy risk. The precise underlying factors responsible for this increased risk will require further study, and may include the chronic stress of hemolysis, an inflammatory bone marrow microenvironment, stressful in vitro cell manipulation, and exposure to cytotoxic conditioning regimens. We believe that the rapid reporting of these results will assist in the ongoing search for the causes of myeloid malignancies in SCD and help define ways to reduce this risk, which is particularly relevant to current and future genetic therapies for this disease.

## 2. Methods

### 2.1. WGS Samples

30 studies from the TOPMed consortium were compiled together into a single dataset. The studies were the following: Amish, ARIC, BAGS, BioMe, CARDIA, CFS, CHS, COPDGene, CRA, DHS, FHS, GALAII, GeneSTAR, GenSalt, GOLDN, HCHS SOL, HyperGEN, JHS, MESA, MLOF, OMG SCD, REDS-III Brazil, SAFS, Samoan, SAPPHIRE asthma, SARP, THRV, VU AF, walk PHaSST, WHI. Together, the studies had 71,100 individuals unaffected by SCD and 3,090 individuals affected by SCD, with a mean of 2,982 individuals per cohort.

### 2.2. Somatic Mutation Calling

Somatic mutations were called from WGS samples using GATK-Mutect2 in combination with GATK-Mutect2-PON, as we described previously[14]. Analysis was performed using publicly available methods in Workflow Description Language (WDL) on the Broad Institute’s Terra Platform, and BAM files were remapped and harmonized through a previously described unified protocol. Single nucleotide polymorphisms (SNPs) and short indels were jointly discovered and genotyped across the TOPMed samples using the GotCloud pipeline. An SVM filter was trained to discriminate between true variants and low-quality sites. Sample quality was assessed through pedigree errors, contamination estimates, and concordance between self-reported sex and genotype-inferred sex. Variants were annotated using snpEff 4.3. Putative somatic SNPs and short indels were called with GATK Mutect2 (https://software.broadinstitute.org/gatk). In brief, Mutect2 searches for sites where there is evidence for variation and then performs local reassembly. It uses an external reference of recurrent sequencing artefacts termed a ‘panel of normal samples’ to filter out these sites, and calls variants at sites where there is evidence for somatic variation. The panel of normal samples used for our study included 100 randomly selected individuals under the age of 40 years. Absence of a hotspot CH mutation was verified before inclusion in the panel of normal set. An external reference of germline variants was provided to filter out likely germline calls. We deployed this variant calling process on Google Cloud using Cromwell (https://github.com/broadinstitute/cromwell). The caller was run individually for each sample with the same settings. The Cromwell WDL configuration config files for the run conditions found in this manuscript can be found on the Sankaran lab github (https://github.com/sankaranlab/scd-chip).

### 2.3. Sickle Trait Identification

Individuals were genotyped and called using the GotCloud/vt pipeline in Freeze 8 of TOPMed. Bcftools was used to extract the rs334 locus. Any individual carrying a heterozygous call (T/A) at that locus was determined as a sickle trait carrier.

### 2.4. CH Prevalence Modeling

Only somatic variants that were present in a single individual within all cohorts were used for prevalence modeling. To derive effect estimates, a binomial logistic regression model was then fit to the data which was annotated for the presence or absence of a clone indicative of CH. Models were then adjusted, where indicated, for age, age^2^, sex, study, sickle cell genotype (hemoglobin genotype driving the SCD), hydroxyurea use, and the first 10 principal components. At all instances in the text, ORs are adjusted for at least age and age^2^. Graphically, in most cases, generalized additive models were used to represent patterns across age, with shaded regions corresponding to 95% confidence intervals. Model assumptions were checked in all cases. To incorporate HU treatment, individuals were classified as either undergoing HU treatment or not; treatment was not treated as a continuous variable. To build ideally matched control cohorts for individuals with SCD, the first 10 PCs and age were used to select the 20 most closely matching individuals from the entire unaffected cohort for each individual with SCD, using a nearest-neighbour approach on propensity scores, exact matching was performed on sex. From this analysis, smaller disease-specific cohorts (DHS, ECLIPSE, VU AF, CHS, HVH, GOLDN, Mayo VTE, EO-COPD, IPF, ECLIPSE, ARIC, FHS) were excluded. Matching quality was evaluated by Love plots, with a threshold of 0.1 for absolute mean differences as well as visual inspection of each covariate against propensity score, separated by SCD diagnosis. Subsequently, weightings from this matching were used in binomial logistic regression analysis, pair membership was used to ensure estimated effects and variance were cluster robust. This constructed control cohort was then used as a comparison for CH prevalence as matched controls.

### 2.5. CH Gene Ranking

To rank the most commonly mutated genes, singleton variants were filtered to those that are considered to be indicative of the presence of CH. Genes were then ranked by the number of somatic variants they contained across all cohorts.

### 2.6. Power Calculations

Power calculations were performed using all individuals 70 years of age and younger without SCD as the control population, and using all individuals 70 years of age and younger with SCD as the test population. Power cutoffs were binned at 5% thresholds, and cohort SCD cohort numbers were then used to calculate the maximum bin that powered at a particular fold difference in CH. Given that young individuals are unlikely to exhibit CH, we disregard these samples when calculating required power, and require that individuals must be equal to or above the age of 40 years old in order to have a reasonable chance of containing detectable somatic expansions. Within this age range, all individuals combined across cohorts without SCD had a 3.76% rate of CH. The experimental population with SCD equal to or above the age of 40 contained 478 individuals. The probability of Type I error (alpha) used was 0.05. In order to detect a 2X or 7.52% rate of CH within the experimental SCD population with 95% power, a minimum of 460 individuals are required. To detect a 1.75X rate of CH with 80% power, a minimum of 425 individuals are required.

### 2.7. SBS Mutation Spectra

To measure relative rates of SBS in each individual, somatic mutations were first binned by individual and annotated by SCD status. Using hg38 as the reference genome, the neighboring upstream and downstream bases were associated with each somatic substitution mutation to define the trinucleotide single base substitution for each change. SigProfilerMatrixGenerator was then used to concatenate together each of these trinucleotide changes and compile a full matrix containing all individuals used in this study [20]. The relative occurrences of these trinucleotide changes were then calculated per individual and, plotted across all individuals with standard deviation as the estimation of error.

## Data Availability

Individual WGS data for TOPMed whole genomes, individual-level harmonized phenotypes, harmonized germline variant call sets, and the CH somatic variant call sets are available through restricted access via the dbGaP. Accession numbers for these datasets have been described in our prior study14, except for REDS-III_Brazil (phs001468), OMG_SCD (phs001608), and walk_PHaSST (phs001514).

## 3. Data Availability

Individual WGS data for TOPMed whole genomes, individual-level harmonized phenotypes, harmonized germline variant call sets, and the CH somatic variant call sets are available through restricted access via the dbGaP. Accession numbers for these datasets have been described in our prior study [14], except for REDS-III Brazil (phs001468), OMG SCD (phs001608), and walk PHaSST (phs001514).

## 4. Acknowledgments

We are grateful to members of the Sankaran laboratory and numerous colleagues for valuable comments and suggestions. This work was supported by the New York Stem Cell Foundation (V.G.S.), a gift from the Lodish Family to Boston Children’s Hospital (V.G.S.), the Burroughs Wellcome Fund Career Award (A.G.B.), and National Institutes of Health Grants R01 DK103794 (V.G.S.), R01 HL146500 (V.G.S.), and DP5 OD029586 (A.G.B.). L.A.L. received support from National Institute of Health Grant T32 HL066987. V.G.S. is a New York Stem Cell-Robertson Investigator.

Molecular data for the TOPMed program was supported by the National Heart, Lung and Blood Institute (NHLBI). WGS for “NHLBI TOPMed: DHS” (phs001412) was performed at the Broad Institute Genomics Platform (HHSN268201500014C). WGS for “NHLBI TOPMed: Samoan” (phs000972) was performed at the Northwest Genomics Center (HHSN268201100037C) and the New York Genome Center Genomics (HHSN268201500016C). WGS for “NHLBI TOPMed: walk PHaSST” (phs001514) was performed at the Baylor Genomics Center (HHSN268201500015C). All other cohorts have been previously analyzed and acknowledged in a prior study on CH [14]. Core support including centralized genomic read mapping and genotype calling, along with variant quality metrics and filtering were provided by the TOPMed Informatics Research Center (3R01HL-117626-02S1; contract HHSN268201800002I). Core support including phenotype harmonization, data management, sample-identity QC, and general program coordination were provided by the TOPMed Data Coordinating Center (R01HL-120393; U01HL-120393; contract HHSN268201800001I). We gratefully acknowledge the studies and participants who provided biological samples and data for TOPMed.

## 4.1. Walk-PHaSST Acknowledgement

We thank Dr. Mark Gladwin and the investigators of the Walk-PHaSST study and the patients who participated in the study. We also thanks the Walk-PHaSST clinical site team: Albert Einstein College of Medicine: Jane Little and Verlene Davis; Columbia University: Robyn Barst, Erika Rosenzweig, Margaret Lee and Daniela Brady; UCSF Benioff Children’s Hospital Oakland: Claudia Morris, Ward Hagar, Lisa Lavrisha, Howard Rosenfeld, and Elliott Vichinsky; Children’s Hospital of Pittsburgh of UPMC: Regina McCollum; Hammersmith Hospital, London: Sally Davies, Gaia Mahalingam, Sharon Meehan, Ofelia Lebanto, and Ines Cabrita; Howard University: Victor Gordeuk, Oswaldo Castro, Onyinye Onyekwere, Vandana Sachdev, Alvin Thomas, Gladys Onojobi, Sharmin Diaz, Margaret Fadojutimi-Akinsiku, and Randa Aladdin; Johns Hopkins University: Reda Girgis, Sophie Lanzkron and Durrant Barasa; NHLBI: Mark Gladwin, Greg Kato, James Taylor, Wynona Coles, Catherine Seamon, Mary Hall, Amy Chi, Cynthia Brenneman, Wen Li, and Erin Smith; University of Colorado: Kathryn Hassell, David Badesch, Deb McCollister and Julie McAfee; University of Illinois at Chicago: Dean Schraufnagel, Robert Molokie, George Kondos, Patricia Cole-Saffold, and Lani Krauz; National Heart & Lung Institute, Imperial College London: Simon Gibbs. Thanks also to the data coordination center team from Rho, Inc.: Nancy Yovetich, Rob Woolson, Jamie Spencer, Christopher Woods, Karen Kesler, Vickie Coble, and Ronald W. Helms. We also thank Dr. Yingze Zhang for directing the Walk-PHasst repository and Dr. Mehdi Nouraie for maintaining the Walk-PHaSST database and Dr. Jonathan Goldsmith as a NIH program director for this study. Special thanks to the volunteers who participated in the Walk-PHaSST study. This project was funded with federal funds from the NHLBI, NIH, Department of Health and Human Services, under contract HHSN268200617182C. This study is registered at www.clinicaltrials.gov as NCT00492531. Detailed description of the study was published in Blood, 2011 118:855-864, Machado et al “Hospitalization for pain in patients with sickle cell disease treated with sildenafil for elevated TRV and low exercise capacity”.

## 4.2. Diabetes Heart Study (DHS) Acknowledgements

This work was supported by R01 HL92301, R01 HL67348, R01 NS058700, R01 AR48797, R01 DK071891, R01 AG058921, the General Clinical Research Center of the Wake Forest University School of Medicine (M01 RR07122, F32 HL085989), the American Diabetes Association, and a pilot grant from the Claude Pepper Older Americans Independence Center of Wake Forest University Health Sciences (P60 AG10484).

## 4.3. Samoan Adiposity Study (Samoan) Acknowledgements

Date collection was funded by NIH grant R01-HL093093. We thank the Samoan participants of the study and local village authorities. We acknowledge the support of the Samoan Ministry of Health and the Samoa Bureau of Statistics for their support of this research.

## 4.4. Author Contributions

L.A.L., L.D.C., J.S.W., A.G.B., and V.G.S. conceived and designed the study, and wrote the manuscript with input from all authors. L.A.L., L.D.C., and J.S.W. analyzed data. Y.Z., S.M.N., M.T.G., M.E.G., A.A-K., M.J.T., B.C., S.K., C.L.D., E.C.S., P.L., A.B.C-P., C.M., A.P.R, G.R.A., D.A.W., and P.N. contributed clinical data or analytic advice. A.G.B. and V.G.S. supervised the study. Competing Interests G.R.A. is an employee of Regeneron Pharmaceuticals and owns stock and stock options for Regeneron Pharmaceuticals. P.N. reports research grants from Amgen, Apple and Boston Scientific, and is a scientific advisor to Apple and Blackstone Life Sciences, all unrelated to the present work.

